# The health and economic impacts of a heat wave: a scenario-based risk assessment

**DOI:** 10.64898/2026.06.29.26356451

**Authors:** Alison Kelly, Richard Bruns, Hannah Goodtree, Amanda Mui, Crystal Watson

**Affiliations:** SHIELD Initiative, Center for Health Security, Johns Hopkins Bloomberg School of Public Health; Center for Outbreak Response Innovation, Johns Hopkins, Bloomberg School of Public Health, Baltimore, Maryland, United States of America; Max Welfare; Biosecurity and Pandemic Policy Center, The Scowcroft Institute of International Affairs at the Bush School of Government & Public Service, Texas A&M University, Washington, DC, United States of America

## Abstract

The impact of weather on the health of Americans and the American health system is substantial. Using available health and economic data, we developed a data-driven scenario that describes a compounded heat emergency in an archetypal community in the United States. We then characterize the potential human and economic costs of such a heat emergency to demonstrate the widespread impact on health outcomes, health systems, and society.

## Introduction

Extreme heat is the most common cause of weather-related fatalities (1). Exposure to several days of unusually hot weather (compared to the historical averages) is generally considered a heat wave. Heat waves have the potential to cause large numbers of deaths, with recorded impacts ranging from 700 in Chicago in 1995 to over 70,000 in Europe in 2003 (1–3); the true numbers are likely higher, as heat-related deaths are often underreported (2). Heat waves pose particular challenges for preparedness and response, as they are less visible than other types of natural hazards, the health consequences can have a slower or lagged onset, and heat-related illnesses are not notifiable conditions and are subject to misclassification.

Primary heat-related illnesses include heat exhaustion and heat stroke, while secondary illnesses include exacerbations of chronic illnesses such as cardiovascular disease, respiratory disease, kidney disease, and mental illness (3). Heat can also contribute to other adverse outcomes, such as injuries from accidents or violence (4,5). This means that health impacts of heat will be experienced across the health system, including emergency medical services, outpatient clinics, and hospital systems.

The costs of heat-related illnesses are also high. In 2020, excess Medicaid spending on heat-related illnesses, including heat stress, electrolyte imbalance, acute myocardial infarction, and acute renal failure, totaled nearly $200 million (6). Using data from Virginia, extrapolated nationally, modelers have estimated that heat-related or heat-adjacent illnesses result in approximately $1 billion in health care costs in the United States annually (7,8).

Fortunately, most heat-related illnesses and deaths are preventable through advanced preparation by governmental agencies at the federal, state, and local levels in collaboration with health systems and communities. Effective management of a heat emergency requires multi-sector emergency coordination. Some communities, particularly those that have experienced a recent heat emergency, have implemented plans to address this threat (9,10). However, it is increasingly apparent that virtually all communities in the United States are at risk for heat-related health emergencies, which can occur at lower temperatures than commonly understood, and that the wide range of potential impacts is not well understood. Furthermore, heat events have a high probability of being compounded events, meaning that they occur contemporaneously with other events such as wildfires, hurricanes, mass gatherings or infectious disease outbreaks. Power outages associated with severe weather are also increasingly common across the United States. Between 2018 and 2020, in 1,657 counties with reliable data, 50.6% of counties (mostly in the Southeast) experienced a power outage of greater than 8 hours in association with an anomalous heat event; when a compounded event with both anomalous heat and anomalous precipitation, 8-plus-hour power outages occurred for a total of 1,155 county-days across 40 states (10). For these reasons, virtually every community could benefit from considering the risks and costs associated with a heat emergency.

A recommended first step in developing a heat action plan might include a scenario-based qualitative risk assessment based on specific local conditions. The hypothetical example presented here offers one such data-driven scenario to consider. This scenario is intended as a planning-support tool for preparedness discussions and should not be interpreted as a predictive forecast.

### Heat Wave Scenario Description

This scenario is intended as a planning-support tool for preparedness discussions and should not be interpreted as a predictive forecast. A prolonged heat wave with daytime highs of 96–104°F (36–40°C) and nighttime lows >70°F hits a state in late July. During this same 5-day period, the state is hosting two large spectator events at outdoor facilities. Each event is likely to draw local and out-of-town spectators, volunteers, media, and staff into the facilities and the surrounding neighborhoods. Each mass gathering is expected to draw 20,000-30,000 participants; one event is a single afternoon sporting event, while the other is a weekend-long music festival with activities occurring throughout the day.

On day 3 of the heat wave, the power grid in the area reaches peak demand, and there are rolling brownouts affecting approximately 250,000 customers for about 24 hours.

The urban area is the most impacted. Many residents are left without air conditioning or power for home medical devices, and hospitals are forced to delay or cancel some elective procedures while they are on backup power.

Meanwhile, the two events continue, with many activities running on generator power. Some vendors are affected, and food preparation becomes more difficult. At the two mass gatherings, thousands of people seek treatment for heat-related illness, and some of those are transported to area hospitals. In addition, the events generate several food-related outbreaks because of the hot conditions and rolling brownouts, which make some food unsafe for consumption.

In nearby rural areas, agricultural workers are also hard hit, with heat exposures driving increased illness and death, which will also drive future worker compensation costs.

### Probable health care system impacts

The health care system is impacted by both the increased number of visitors to the region and the unusually high temperatures. This combination of mass gatherings plus the heat will strain emergency medical services, urgent care, and hospital systems.

Primary heat illnesses and secondary exacerbations of chronic illnesses will drive an increase in patients needing care across the region. Older adults are most vulnerable to heat illness and are also disproportionately affected by chronic illnesses like heart disease, asthma, and other chronic respiratory diseases that can be worsened by excess heat.

Assuming adequate preparation of the facilities, many of the health concerns associated with the mass gathering events will be handled by on-site first-aid, medical and emergency service providers. However, urgent care and emergency departments near the venues will experience upticks in low acuity patients presenting with e.g., dehydration, alcohol-related injuries and conditions, minor injuries, as well as a limited number of higher acuity presentations, such as cardiac events.

With the rolling brownouts, there will also be impacts on patients who need dialysis and patients who rely on electric-powered medical devices. Some of these patients will need acute care due to their device failures, and some will require transportation from emergency medical services. Moreover, there will be long-term health effects from delayed care and cancelled elective procedures.

This scenario considers a moderate heat event, with temperatures that would be considered high for some regions of the country. Impacts can occur in milder scenarios and would be exacerbated by more extreme scenarios. Regional differences in normal temperatures and ability to respond when the weather is hotter than normal (e.g., cooling availability in homes and businesses) affect what is considered a heat wave and its impacts.

### Heat Wave Scenario Assumptions

Assumptions are derived from historical examples and reports from peer-reviewed and grey literature sources.

### Population and Epidemiologic Assumptions

The population assumptions are based on data for the archetypal location of Baltimore/Maryland. Baltimore City and surrounding Maryland counties were chosen as the archetype for several reasons:

- Population and health data were transparent and accessible.
- Like many other cities/states in the US, the population of Baltimore/MD is reasonably acclimated to heat, but not to an anomalous heat event that is prolonged.
- Like other cities, Baltimore has documented urban heat island intensity, aging housing, and vulnerable populations with limited access to AC in their homes.
- Baltimore has a large, elderly, minority population with high burdens of heat-exacerbated comorbidities, such as hypertension, chronic kidney disease, and heart failure. Something that urban and rural populations across the country share.
- Maryland has rural populations with large numbers of agricultural workers and gaps in EMS service and healthcare access. This is echoed in many other US rural communities.
- Like most other states, Maryland’s healthcare systems are operating under strain, largely at or near capacity on a daily basis.

While the population assumptions are based on Baltimore, this scenario is an archetype and could also apply to similarly sized metropolitan areas across the US, such as Charlotte, Orlando, Saint Louis or San Antonio. The scenario does not presume any specific capabilities of the public health or health care system in Baltimore and should not be considered to be predictive of an event occurring in Baltimore.

Population and Impact Assumptions
| Category of impacts | Population exposed at baseline | Scenario assumptions |
| --- | --- | --- |
| **Heat-related excess deaths** | Metro area population of 2.9 million plus 100,000 mass gathering participants (11,12) | 4–15 deaths per 100,000 population, estimated from past events (13,14). |
| **EMS dispatches above baseline** | Baseline of 1,500 EMS calls/day | Past events show a 30–80% increase in EMS dispatches; a 50% mid-range increase was selected (15). |
| **Food-borne illness cases** | Participants in the mass gatherings | Attack rate at outdoor events with temperature-control failures is 2–8%. Scenario assumes 45,000 food consumers and a 3% attack rate (16). |
| **Heat-related medical presentations at the outdoor events \*\*** | 100,000 total attendees across the 2 events | 15–30 per 1,000 per day heat-related illnesses for the events. |
| **Heat-related ED visits (general population and agricultural workers)** | Metro area population of 2.9 million plus 100,000 mass gathering participants (11,12) and 17,500 agricultural workers | Baseline ED visit rate over 5 days is 5.2 per 100,000; a heat wave increases this 4-fold. Assumes 7–10% of the 2,000 event participants presenting with heat-related symptoms require EMS transport (8,17). Agricultural worker rate: 15–40 per 1,000 (18–21). |
| **Hospitalizations\*\*\*** | From ED admissions, food-borne disease admissions, and direct admissions for medical equipment failures or chronic disease exacerbations | See category details below. Includes direct admissions of patients who experienced home medical device failure (22,23). |
| **Heat-related illnesses (general population)** | Metro area population of 2.9 million plus 100,000 mass gathering participants (11,12) | ~22% of heat-related ED visits require hospitalization in the general population; assumed higher for mass gatherings at 30–45% (8,24). |
| **Food-borne illness hospitalizations** | Participants in the mass gatherings | About 1.9% of Salmonella cases result in hospitalization (16). |
| **Home medical equipment failures** | 10,000 home device users in the metro area, estimated from national prevalence data | 10% of home medical device users may require emergency intervention during a 24-hour power outage (22,23). Dialysis and other equipment failure costs are calculated separately. |
| **Dialysis patients unable to access treatment** | 6,500 dialysis patients in the metro area, estimated from national prevalence data | 25% of dialysis patients are unable to access treatment and require emergency intervention during the power outage (22,23). |
| **Cardiovascular and respiratory disease exacerbations \*** | Metro area population of 2.9 million (11,12) | Heat emergencies bring a 3–14% increase in healthcare utilization; ~40% is CV or respiratory. Yields an estimated 3,000 |
|  |  | CV/respiratory exacerbations over the 5 days (25,26). |
| **Workers' compensation for agricultural workers (22–25)** | 17,500 workers | 15–40 per 1,000 heat-related illness rate in agricultural workers (18–21). |
| **Elective procedures cancelled** | 4 metropolitan area hospitals | Assumes ~50 elective procedures/day per hospital (200 total) and that ~19% are cancelled (25,27). |
| *\* Not nationally costed; modeled as excess hospitalization.*<br>*\*\* Expert-opinion estimate for first aid at a notional outdoor event, not substantiated by a specific source.*<br>*\*\*\* Hospitalizations due to home medical equipment failures, chronic disease exacerbations, and dialysis treatment access difficulties are not included in emergency department visits in this model due to an assumption that they would be quickly assessed in the ED and admitted.* |  |  |

### Modeled Impacts and Costs for the Scenario

The analysis included a Monte Carlo simulation. Most calculation inputs were in the form of probability distributions rather than point estimates. Results are reported as ranges, which represent the 90% confidence interval; in other words, the 5th and 95th percentile values from the Monte Carlo simulation (90% CI). All costs in USD millions [90% CI]. Figures are rounded to two significant figures for readability and to avoid false precision.

| Cost Category | Number of events | Medical system costs | Other social costs | Monetized DALY loss | Total costs |
| --- | --- | --- | --- | --- | --- |
| **Heat Wave Associated Excess Deaths** |  |  |  |  |  |
| Heat-related excess deaths | 300<br>[190, 410] | 0.36<br>[0.23, 0.49] | 3.5<br>[1.8, 5.8] | 700<br>[170, 1530] | 710<br>[180, 1540] |
| **Heat Wave Associated General Costs** |  |  |  |  |  |
| EMS dispatches above baseline | 4000<br>[2800, 5400] | 4.8<br>[3.3, 6.4] | 2.2<br>[1.3, 3.3] | 1.1<br>[0.6, 1.7] | 8.1<br>[5.5, 11.1] |
| **Mass Gathering Event General Costs** |  |  |  |  |  |
| Food-borne illness cases from mass gathering events | 1900<br>[1100, 3200] | 0.41<br>[0.23, 0.67] | 1.1<br>[0.5, 1.9] | 6.5<br>[3.1, 11.1] | 7.9<br>4.1, 13.4] |
| Heat-related medical presentations at mass gatherings | 2200<br>[1600, 2900] | 0.88<br>[0.63, 1.15] | n/a | 0.3<br>[0.17, 0.45] | 1.2<br>[0.8, 1.6] |

| ED Visits (combined heat wave and mass gathering events) |  |  |  |  |  |
| --- | --- | --- | --- | --- | --- |
| Heat-related ED visits (general population) | 910 (220 from mass gathering)<br>[670, 1180] | 0.69<br>[0.51, 0.89] | 0.51<br>[0.3, 0.75] | 0.25<br>[0.15, 0.36] | 1.4<br>[1, 1.9] |
| Heat-related ED visits (workers) | 470 (180 from mass gathering)<br>[310, 650] | 0.36<br>[0.24, 0.49] | 5.7<br>[3.8, 7.8] | 0.13<br>[0.07, 0.2] | 6.2<br>[4.1, 8.4] |
| Hospitalizations (combined heat wave and mass gathering events) |  |  |  |  |  |
| Heat-related illnesses (general population) | 160 (85 from mass gathering)<br>[100, 220] | 2.4<br>[1.6, 3.33] | 0.44<br>[0.24, 0.69] | 4.6<br>[1.3, 10.1] | 7.4<br>[3.7, 13.5] |
| Heat-related illnesses (workers) | 25 (10 from mass gathering)<br>[16, 36] | 0.37<br>[2.1, 5.5] | 0.069<br>[0.038, 0.11] | 0.72<br>[0.18, 1.53] | 1.2<br>[0.6, 2.1] |
| Event-only-related hospitalizations |  |  |  |  |  |
| Food-borne illnesses hospitalization from events | 37<br>[21, 60] | 0.21<br>[0.12, 0.35] | 0.041<br>[0.019, 0.073] | 25<br>[10, 49] | 26<br>[10, 49] |
| Heat-wave-only related hospitalizations |  |  |  |  |  |
| Home dialysis failures | 1600<br>[1300, 2000] | 24<br>[19, 30] | 4.5<br>[2.8, 6.4] | 47<br>[13, 96] | 76<br>[39, 127] |
| Other home medical device failures | 730<br>[500, 1020] | 11<br>[7, 15] | 2<br>[1.2, 3.1] | 21<br>[5, 46] | 34<br>[17, 61] |
| CV/respiratory disease exacerbations | 250<br>[140, 370] | 3.7<br>[2.1, 5.5] | 0.69<br>[0.34, 1.15] | 25<br>[10, 48] | 29<br>[12, 54] |
| Elective procedures canceled | 210<br>[160, 280] | 0.011<br>[0.008, 0.014] | 0.021<br>[0.01, 0.036] | 14<br>[3, 34] | 14<br>[3, 34] |
| **Total Heatwave Costs** |  | **48**<br>**[40, 56]** | **19**<br>**[15, 25]** | **810**<br>**[260, 1640]** | **870**<br>**[330, 1710]** |
| **Total Heatwave and Event Costs** |  | **51**<br>**[42, 59]** | **21**<br>**[16, 27]** | **850**<br>**[290, 1690]** | **920**<br>**[360, 1760]** |

- **State Impact:** Under a modeled scenario representing today’s levels of public health preparedness, a significant heat wave – a period of unusually hot weather lasting five days – in one state could cause **300** deaths, **1200** heat-related emergency department visits, and **2800** preventable hospitalizations. Those impacts may generate approximately **$870 million** in total costs, including **$48 million** in medical costs, **$19 million** in other social costs and **$810 million** in disrupted livelihoods, lost wages, and diminished well-being.
- **National Impact:** Given that the United States Environmental Protection Agency (EPA) estimates that the U.S. can expect roughly six heat waves each year across its largest metropolitan areas, findings from the modeling analysis suggest total costs for these heat waves could approach more than **$5.22 billion** annually nationwide.
- **Under a more extreme, though still likely, modeled scenario** where two outdoor spectator events are also taking place during the heat wave, one state could experience **$920 million** in total costs, **300** deaths, **1**,**380** heat-related emergency department visits, and **2**,**900** preventable hospitalizations. Those impacts may generate approximately **$920 million** in total costs, including **$51 million** in medical costs, **$20 million** in other social costs and **$850 million** in disrupted livelihoods, lost wages, and diminished well-being. Across the 6 potential heat waves for the year under this scenario, total costs for these heat waves could approach more than **$5.52 billion** annually nationwide.

### Limitations of the scenario and economic model

This is one possible scenario, which was developed to examine the public health, healthcare, and economic impacts of heat-related events. This scenario is fictional and is not predictive of future events. The scenario is grounded in real-world data to the extent available and utilizes past events to guide assumptions and the scenario construct. However, as this is only one scenario, it cannot fully represent possible future events.

The quality and availability of the data used to support the scenario’s assumptions are variable. Past events provide good data on many aspects of a heat emergency scenario but may not be directly applicable to future events or other geographic areas. There was very limited data available on some categories of health impacts of heat, such as impacts on mental health and violence, pregnancy complications and injuries, which were therefore excluded from the analysis. Some categories of health impacts, such as chronic disease exacerbations and failures of home medical devices, had limited cost data and were therefore modeled in the cost analysis as excess hospitalization. It is assumed that patients hospitalized due to failure of home medical devices, chronic disease exacerbations, or lack of access to dialysis treatment would be quickly assessed and admitted to a hospital, and so are not counted as emergency department visits. This may result in an underestimate of ED visit costs for these categories of patients. Healthcare utilization and cost data were extrapolated from multiple sources across the United States and may not fully account for local/regional differences in utilization patterns and costs.

## Discussion

This scenario demonstrates that a heat wave in a single metropolitan area can have profound health and economic consequences. However, the US Environmental Protection Agency estimates that across the major metropolitan areas of the US, 6 heat waves can be expected annually (28), indicating that the national impact is significantly greater. Additionally, heat-related illnesses and deaths do not occur only during heat waves. The HeatRisk health-based forecast, developed by the National Oceanic and Atmospheric Administration’s National Weather Service and the Centers for Disease Control and Prevention (CDC), is a health-based heat forecast that delivers a 7-day outlook for the risk of heat-related illnesses (29). Because this tool accounts for the unique relationships between heat and health at the local level at different times of the year, with forecasts up to a week in advance, it is an excellent resource to support coordinated planning at the local level. Scenario-based planning to explore the possibilities for anomalous or compounded events, such as the ones presented in this analysis, coupled with the use of evidence-based local heat risk forecasts, offers a path towards the prevention of heat-related health consequences. Several rigorous planning frameworks exist to guide the plan development, including CDC’s Public Health Emergency Preparedness Capabilities (30), the Federal Emergency Management Agency’s National Response Framework (31), healthcare coalition guidance from the Department of Health and Human Services’ Assistant Secretary for Preparedness and Response (32), and the Incident Command System/Unified Command concept (33).

Past emergencies have shown that investments in preparedness and response capabilities can be both lifesaving and cost-saving. Modest reductions in the adverse health outcomes associated with a heat wave can have a major impact. In this scenario, a reduction in the number of adverse outcomes in the range of 10-30% could avoid $210 million [60,480] in total monetized social costs, which includes the cost of 59 [27,100] prevented deaths.

## Conclusions

The public health and healthcare impacts of extreme heat events are underappreciated while also very likely to occur in many locations across the United States. We have seen in past emergencies that preparedness and response plans and capabilities have been lifesaving and cost-saving. This scenario analysis provides a tangible example of the types of health and economic consequences that often accompany extreme heat, and demonstrates how coordinated, multi-sector planning is essential to reducing this burden.

## Data Availability

All data produced in the present work are contained in the manuscript

## Acknowledgements

The authors would like to thank a number of reviewers who provided their expert input, including Kody Kinsley, Dr. Paul Pedersen, Dr. Jeremy Hoffman, and Dr. John Hick.

## Common Health Coalition

Chelsea Cipriano, Rebecca Giglio, Dr. Dave Chokshi, Michael Lanza, Dr. Chris Chen, Juhi Patel, and Arfath Chowdhury.

## Milbank Memorial Fund

Dr. Debra Lubar, Dr. Morgan McDonald, Christine Haran

